# SGLT2 inhibition reduces cardiac surgery-associated acute kidney injury: An open-label randomized study

**DOI:** 10.1101/2024.03.27.24304998

**Authors:** Lars I.P. Snel, Maartina J.P. Oosterom-Eijmael, Elena Rampanelli, Yugeesh R. Lankadeva, Mark P. Plummer, Benedikt Preckel, Jeroen Hermanides, Daniel H. van Raalte, Abraham H. Hulst

## Abstract

**Background:** Cardiac surgery-associated acute kidney injury (CSA-AKI) is a common postoperative complication. Currently, no effective preventative strategies exist to mitigate CSA-AKI. Sodium-glucose transporter-2 (SGLT2) inhibitors reduced acute kidney injury (AKI) incidence in large, randomized placebo-controlled, cardiovascular and kidney outcome trials conducted in patients with chronic kidney disease. We hypothesized that perioperative SGLT2 inhibition could also reduce CSA-AKI.

**Methods:** In this open-label phase IV, randomized, parallel-group, pilot study, adult patients undergoing elective cardiac surgery with cardiopulmonary bypass were randomized to receive the SGLT2 inhibitor, empagliflozin (10 mg; oral), once daily three days prior to surgery and continued to two days after surgery compared with standard-of-care. Biomarkers for acute kidney injury (AKI), including serum and urinary neutrophil gelatinase-associated lipocalin (NGAL), serum and urinary kidney injury molecule-1 (KIM-1), and serum hypoxia-inducible factor-1α (HIF-1α) were measured. Additional outcomes included AKI incidence according to Kidney Disease: Improving Global Outcomes (KDIGO) criteria as well as metabolic parameters, including ketone body concentrations and glycemic control.

**Results:** Between March 2022 and April 2023, 55 patients were included (sex: 73% male, age: 66 ± 10 years, BMI: 28 ± 4 kg/m^2^, empagliflozin n = 25, control n = 30) in the intention-to-treat analysis. Empagliflozin significantly reduced the incidence of AKI (20% vs 66.7%; absolute difference 46.7%, 95% CI, –69.7 – –23.6; P=.001). Following surgery, urinary NGAL, and KIM-1 were found to increase in both arms, whereas a significant increment in serum HIF-1α after surgery was solely observed in the control group. We observed no between-group differences in the incidence of (euglycemic) ketoacidosis or hypoglycemic events.

**Conclusions:** Perioperative SGLT2 inhibition, compared with standard of care, significantly reduced the incidence of CSA-AKI. These findings warrant validation in large-scale, double-blind, placebo-controlled, randomized trials.

**Trial Registry:** https://onderzoekmetmensen.nl/en/trial/26563 Identifier: NL9561

**Clinical perspective:** *What Is New?:*

- In this open-label, randomized, controlled, pilot trial perioperative use of sodium glucose transporter-2 (SGLT2) inhibition with empagliflozin significantly reduced the incidence of acute kidney injury (AKI) by 46.7% (95% CI, –69.7 – –23.6; P=.001) compared to the control group.
- The level of ketone bodies increased significantly during cardiac surgery, however, there was no additional effect of empagliflozin treatment.

*What Are the Clinical Implications?:*

- These results suggest that perioperative treatment with SGLT2 inhibitors might decrease the risk of cardiac surgery-associated (CSA)-AKI.
- These findings warrant validation in large-scale, double-blind, placebo-controlled, randomized trial, which is currently ongoing.

## Introduction

Acute kidney injury (AKI) is a common complication following cardiac surgery, with a reported incidence of up to 70%. (1) Cardiac surgery-associated (CSA)-AKI is associated with postoperative morbidity and mortality, prolonged hospital stay, and increased healthcare costs. (2, 3) In addition, CSA-AKI can have long-term consequences, including the increased risk of developing chronic kidney disease (CKD) and reduced life expectancy compared to patients without CSA-AKI. (4–7)

Despite several attempts to prevent CSA-AKI with a variety of pharmacological interventions, no effective mitigation strategies have been developed to date. (2, 8, 9) Sodium-glucose transporter-2 (SGLT2) inhibitors have been reported to prevent AKI in large cardiovascular and kidney outcome trials in patients with or without type 2 diabetes (T2D). (10–13)

SGLT2 inhibitors reduce proximal sodium and glucose re-uptake, leading to an acute reduction in estimated glomerular filtration rate (eGFR) due to activation of tubuloglomerular feedback. This reduction in eGFR leads to a lesser kidney workload. Lower filtered sodium load can reduce kidney oxygen consumption, while SGLT2-induced improvements in insulin sensitivity and substrate metabolism may further reduce kidney oxidative requirements. (14–16) SGLT2 inhibitors may be protective in settings of renal tissue hypoxia during cardiac surgery, which has been directly reported in clinically relevant large animal models and in humans indirectly by measurement of bladder urinary oxygenation. (17, 18) However, the safety of SGLT2 inhibitors in cardiac surgery should also be considered, given the risk of SGLT2 inhibitor-associated perioperative ketoacidosis caused by the fasted state and surgical stress. (19) This serious side effect has prompted the Food and Drug Administration (FDA) to recommend interruption of SGLT2 inhibitors at least 3 days prior to major surgery. (19, 20) Here, we performed an open-label, randomized, pilot study to evaluate the effects of SGLT2 inhibition on biomarkers of AKI, AKI incidence according to Kidney Disease: Improving Global Outcomes (KDIGO) criteria and incidence of ketoacidosis and hypoglycemia in individuals undergoing cardiac surgery. We hypothesized that perioperative SGLT2 inhibition with empagliflozin would reduce both biomarkers of postoperative AKI and the incidence of AKI.

## Methods

### Trial design

We performed a prospective, single-center, open-label, phase IV, randomized, controlled (standard care), balanced (1:1), pilot study. The study protocol was approved by the medical ethics committee of the Amsterdam UMC and by the Dutch competent authority before the initiation of the trial. The trial protocol was registered at the Dutch Trial Register (https://www.onderzoekmetmensen.nl), number NTR9561. An independent study monitor validated good clinical practice adherence and data collection quality. The study was conducted in compliance with the latest version of the Declaration of Helsinki and Good Clinical Practice guidelines. Neither a data safety management board nor a steering committee was involved in the decision-making of this study. All participants signed informed consent prior to any trial-related activity.

### Study participants

Patients eligible for inclusion were aged between 18 and 90 years and scheduled to undergo elective cardiopulmonary bypass (CPB) assisted cardiac surgery at the Amsterdam University Medical Centre (UMC), a tertiary academic hospital in the Netherlands. Exclusion criteria included prior treatment with SGLT2 inhibitors, type 1 diabetes (T1D), T2D with a body mass index (BMI) below 25 (possibly phenotypically resembling T1D), estimated glomerular filtration rate (eGFR) <30 ml/min, systolic blood pressure below 100 mmHg, a history of ketoacidosis, a known or suspected allergy to the trial product, or individuals who were breastfeeding, intending to become pregnant or not using adequate contraceptive methods.

### Intervention

After providing written informed consent, participants were randomly assigned to the empagliflozin or control group. Patients in the empagliflozin group received a single dose of empagliflozin 10 mg orally (Boehringer Ingelheim International GmbH via Alliance Healthcare Nederland B.V.) every morning, starting from three days prior to surgery, until two days after surgery. The control group received no intervention and received standard perioperative care. We initially intended to start empagliflozin treatment seven days before surgery, however, due to logistical constraints in the planning of surgery, it became evident that this approach was not feasible. Therefore, we adjusted the number of treatments before surgery in the protocol from seven to three days prior to surgery before the recruitment of the first participant.

### Procedures

Patients with T2D had their preoperative glucose management adjusted, if necessary, under the supervision of an endocrinology consultant, according to the treatment algorithm shown in Supplementary Figure 1. At the time of inclusion, on the morning of the day of surgery (before the first incision), and on the first and second morning after surgery (D1 and D2), serum and urine samples were collected to measure neutrophil gelatinase-associated lipocalin (NGAL), kidney injury molecule (KIM)-1, hypoxia-inducible factor 1 alpha (HIF1α) and creatinine. Furthermore, arterial blood gases and ketone levels were collected at four specific time points on the day of surgery: before the start of surgery (T1), at time of start of cardiopulmonary bypass (T2), at the end of cardiopulmonary bypass (T3) and at time of transport to the intensive care unit (ICU) (T4). As part of standard perioperative care, patients with insulin-dependent T2D received a continuous glucose/insulin infusion preoperatively. In case ketoacidosis was suspected based on increased anion gap metabolic acidosis with the presence of ketone bodies, patients were to be treated with a mixed glucose/insulin infusion to suppress ketogenesis and subsequently closely monitored and treated according to local practice. Postoperatively, creatinine levels and urinary output were measured according to usual care (creatinine was measured daily and urinary output hourly until the removal of the urinary catheter upon patients’ discharge from ICU). We also recorded glucose levels as measured during routine care (four times daily during ICU admission) and insulin administrations when prescribed by the treating physicians. Adverse events were collected by reviewing in-hospital electronic health records. The study workflow is summarized in Figure 1.

**Figure 1.**
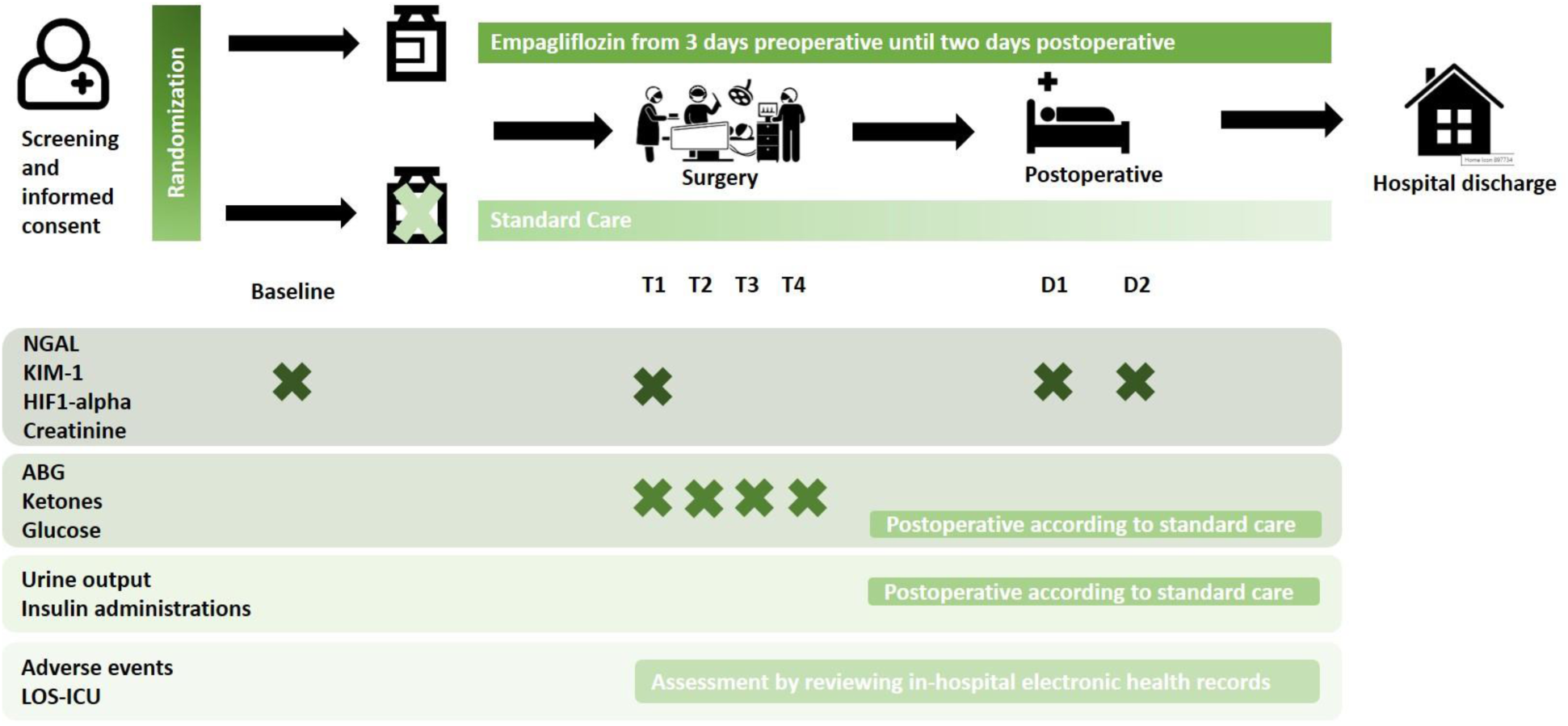
Summary of study workflow.

### Outcomes

The primary outcome was the between-group difference of mean postoperative serum NGAL concentrations measured on postoperative day two. Other outcomes included between-group differences in any of the following measures: AKI incidence rates according to the KDIGO criteria, postoperative urinary NGAL/creatinine ratios, serum KIM-1 concentrations, urinary KIM-1/creatinine ratios, and serum HIF1α levels. Additionally, we calculated the difference in biomarker concentrations between the first and second postoperative days compared to the day of surgery. Metabolic secondary outcomes include the between-group difference of intraoperative ketone levels, the number of ketoacidosis events (defined as a combination of ketonemia >3 mmol/l in combination with a high-anion gap metabolic acidosis), the mean intraoperative glucose level, the mean peak postoperative glucose level measured in the first 48 hours postoperatively, the incidence of hypoglycemia (defined as glucose < 4.0 mmol/L), the incidence of hyperglycemia (defined as glucose > 10.0 mmol/L) and the incidence of insulin administrations. Additionally, we compared the length of stay in the ICU and postoperative in-hospital complications such as mortality, renal replacement therapy, cerebrovascular accident (CVA), cardiac tamponade, gastrointestinal bleeding, anemia requiring transfusion, atrial fibrillation, other cardiac arrhythmia’s, pacemaker implantations, infection of the mediastinum, pneumonia, and delirium.

### ELISA

Serum and urinary concentrations of NGAL and KIM-1 were measured with, respectively, the LEGEND MAX™ Human NGAL (Lipocalin-2) ELISA Kit (Biolegend, cat.N. 443407) and the human KIM1 (TIM-1) ELISA Kit (ABCAM, cat.N. AB235081). Serum HIF-1A levels were detected using the HIF1A Human ELISA Kit (Invitrogen, cat.N. EHIF1A). All measurements were performed after the study had concluded and done according to the manufacturer’s instructions. Urinary levels of NGAL and KIM-1 were normalized for concentrations of creatinine, detected by standard tests at the hospital laboratory of clinical chemistry.

### Sample size

Based on a previous cohort study that found a difference in mean serum NGAL of 120 ng/ml between patients with and without AKI after cardiac surgery, we defined a between-group difference of 120 ng/ml in serum NGAL concentration on postoperative day 2 as a relevant outcome. (21) To detect a difference of 120 ng/ml, with 90% power, alpha at 0.05, we would need 40 patients per group.

### Randomization and blinding

Randomization was performed electronically through the data management system Castor EDC (Ciwit BV, Amsterdam, the Netherlands) using balanced-block randomization, with variable-size random computer-generated blocks of two, four, or six and an allocation ratio of 1:1. By design (open-label study), patients, care providers nor study personnel were blinded to treatment allocation.

### Statistical analyses

We performed all analyses on the intention-to-treat population. When biomarkers were below the detection limit, we assigned the value of zero. Discrete data are presented as counts with percentages per group. Continuous variables are presented as mean ± SD or median (interquartile range), depending on the distribution of the data. Within-group comparisons were conducted using the paired Student t-test. Between-group comparisons were performed using Fisher’s exact test for dichotomous outcomes and unpaired Student’s t-test or Mann-Whitney U-tests for continuous variables, depending on the distribution of the data. Absolute differences between groups are presented with their corresponding 95% confidence intervals (95% CI). Repeated measurements were analyzed with generalized linear mixed models. To compare between the treatment groups, we used the interaction term of the randomization group with time. Additionally, we analyzed postoperative kidney biomarkers by calculating the change in concentration compared to the day of surgery. Statistical analyses were performed using SPSS (IBM version 24; IBM Corp., Armonk, New York).

## Results

Between March 2022 and March 2023, we enrolled 60 patients planned to undergo cardiac surgery, of which 29 patients were included in the empagliflozin group and 31 in the control group (Figure 2). Before the start of the intervention, five patients were excluded (one withdrew consent, two surgeries were cancelled for logistic reasons, and in two cases, the type of surgery was changed, thereby not meeting the inclusion criteria). All remaining patients were included in the intention-to-treat analysis (25 in the empagliflozin group, 30 in the control group). All patients in the intervention group started with empagliflozin three days prior to surgery.

**Figure 2.**
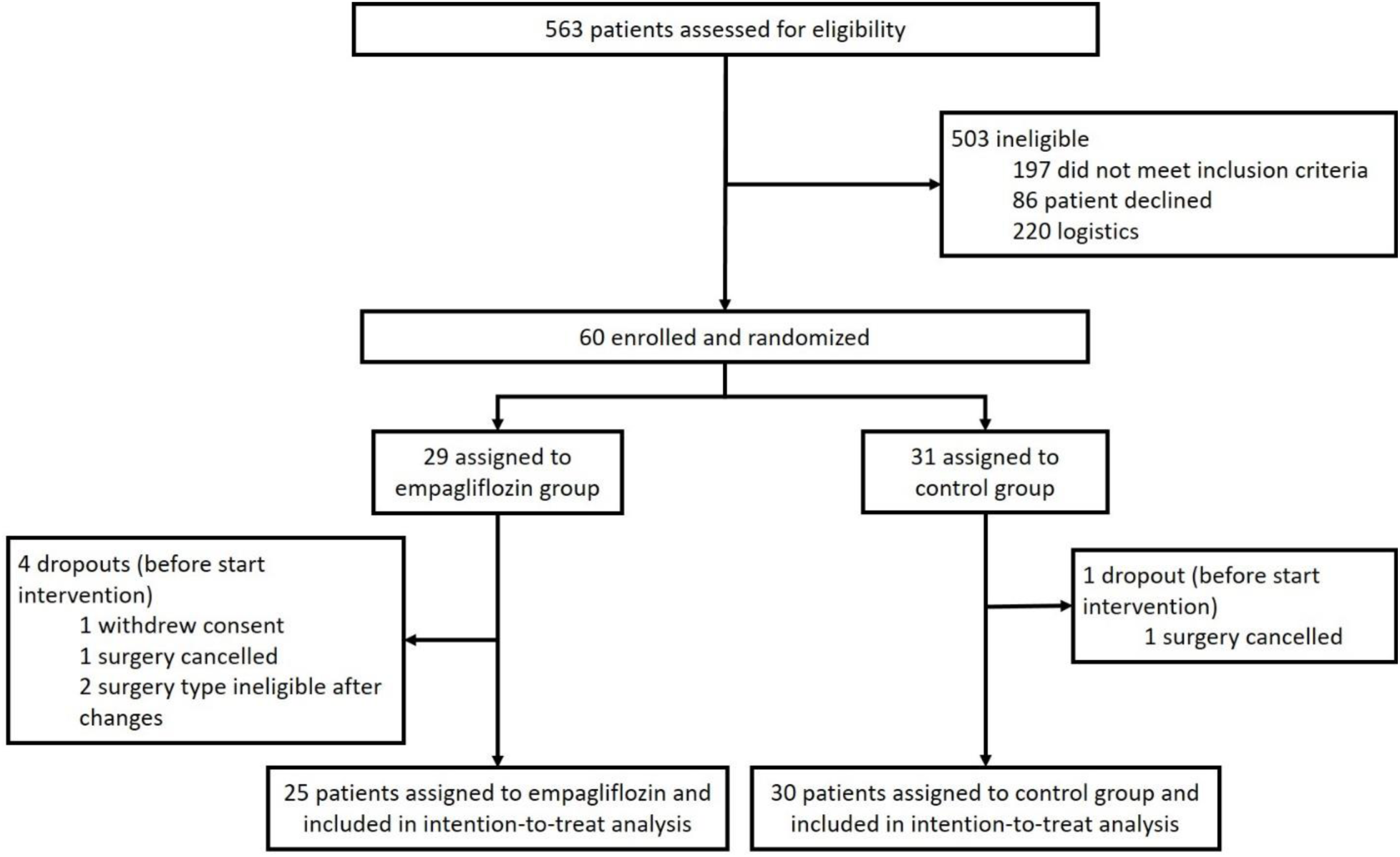
CONSORT flowchart of patients in the study.

Baseline characteristics of all included patients are summarized in Table 1. Overall, patient characteristics were well balanced between groups, except for age, with patients in the empagliflozin group being older. Patients were predominantly male, with relevant overweight, and had normal kidney function at baseline. T2D was present in 5 (9%) patients; none of them were treated with insulin therapy prior to hospital admission.

**Table 1.**
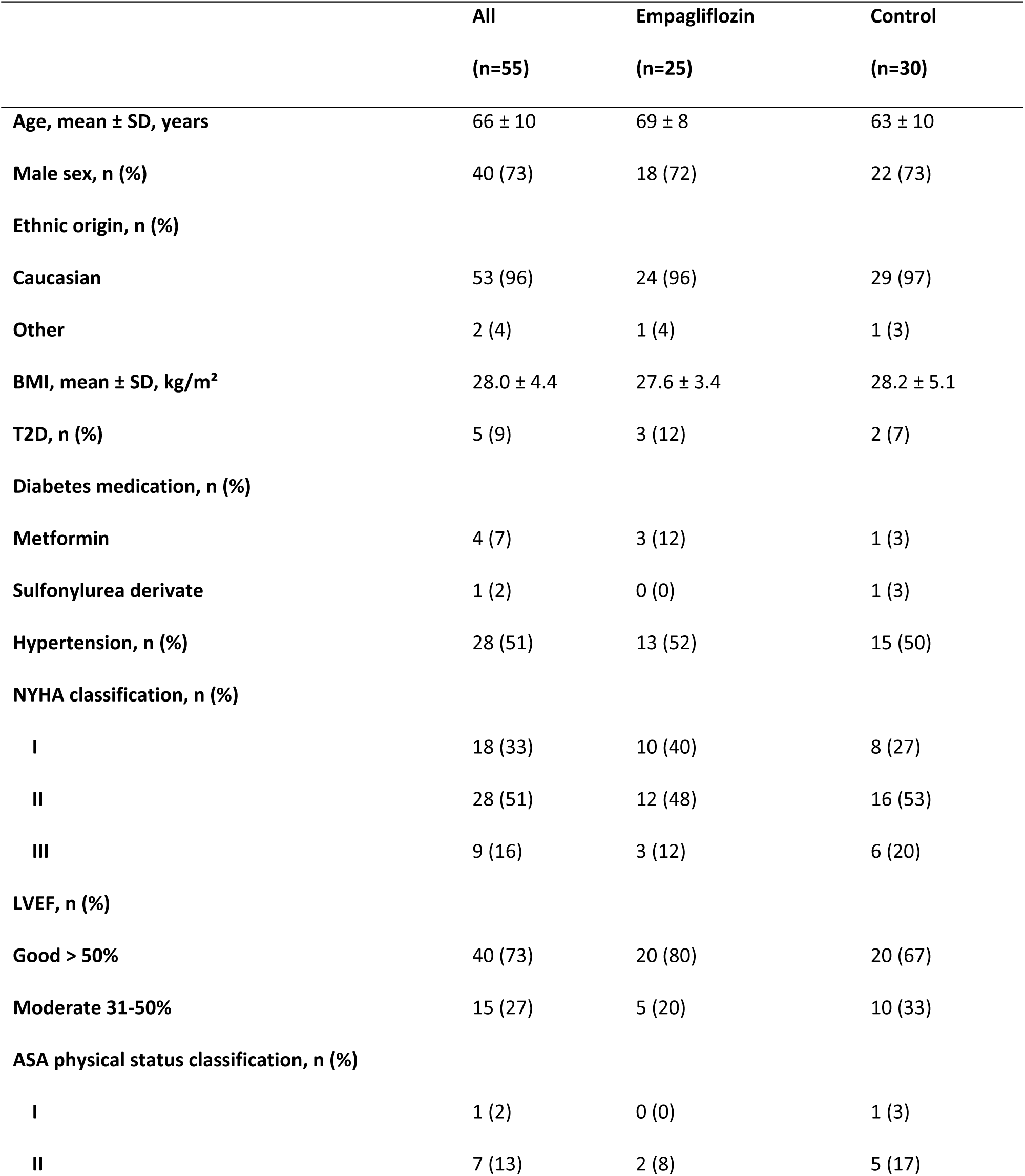
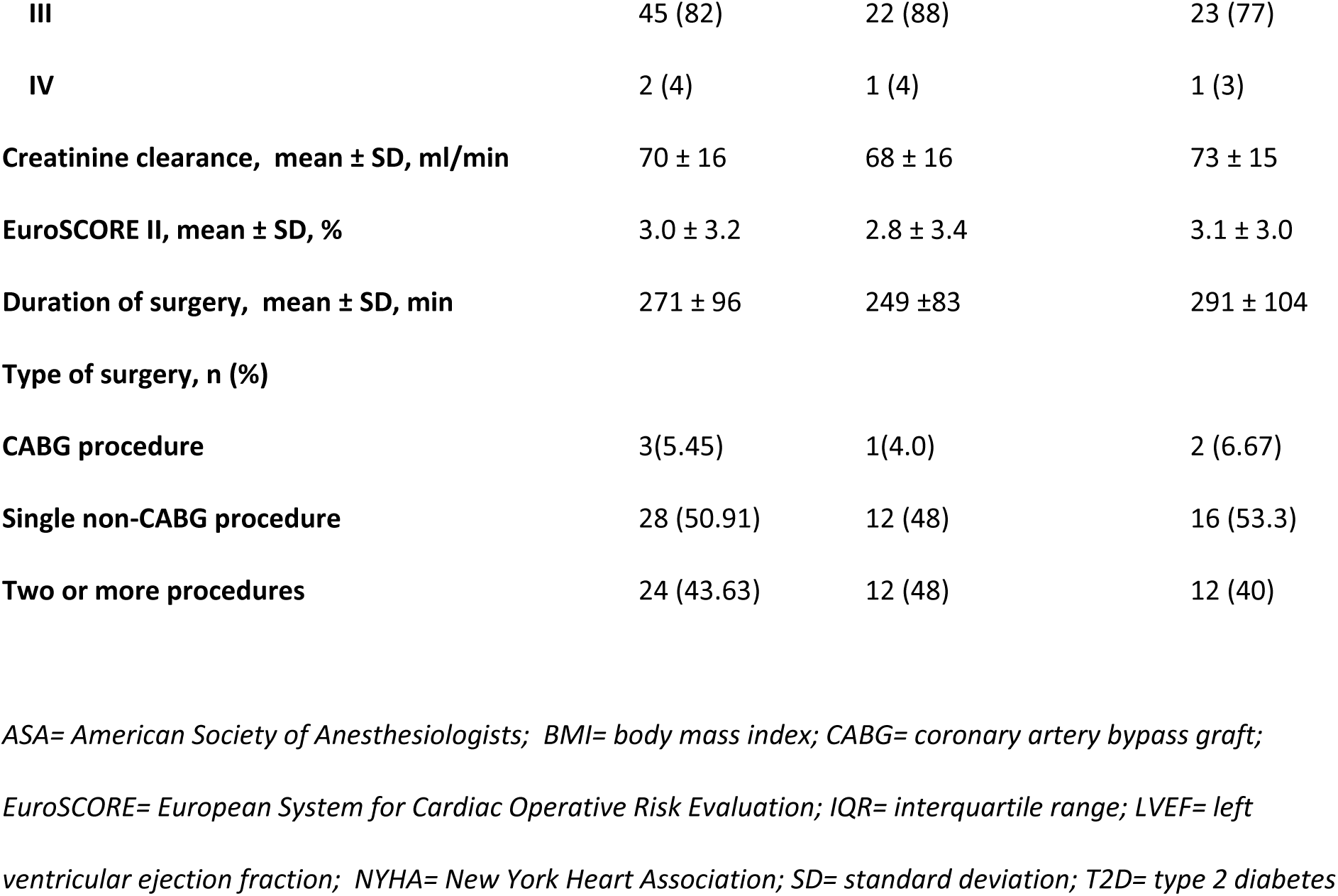
Baseline characteristics of the intention-to-treat population.

### Kidney injury

The development of AKI characterized according to KDIGO criteria was observed in 5/25 (20%) patients in the empagliflozin group and 20/30 (66.7%) patients in the control group, with a difference of 46.7% between groups (95% CI, –69.7 – –23.6; P=.001) (Figure 3). In March 2023, the trial leadership team reviewed this significant between-group disparity in clinical outcomes. Concluding that these findings would require confirmation in a large, blinded and placebo-controlled trial, the trial leadership team ceased patient enrollment in this open-label pilot trial and notified the medical ethics committee of trial termination. Upon completion of data collection, blood and urine samples were sent for analysis.

**Figure 3.**
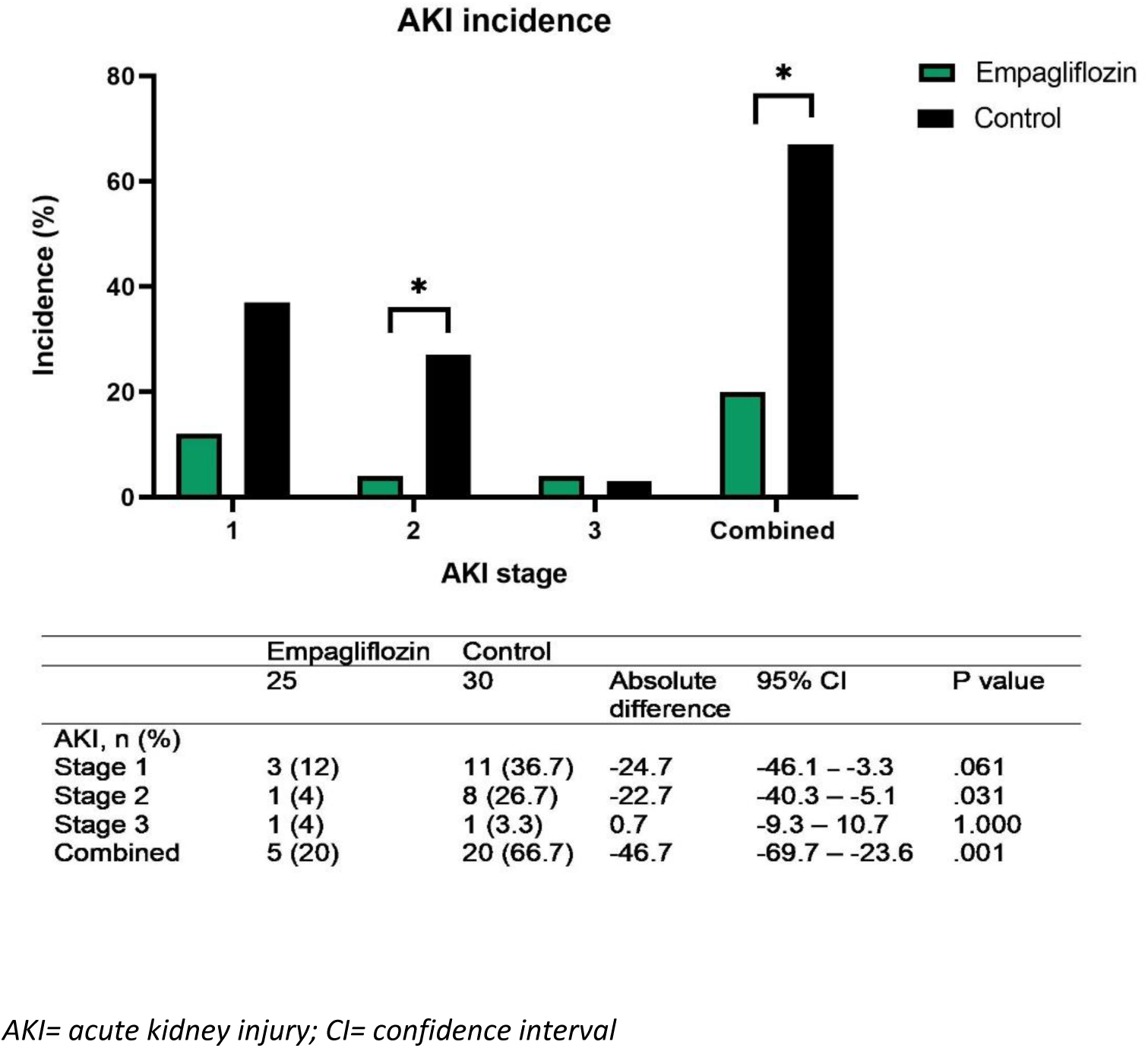
Incidence rates of Acute Kidney Injury (AKI) across various stages compared between the empagliflozin and control group.

### Kidney biomarkers

Serum NGAL concentrations on the second postoperative day did not differ significantly between groups, empagliflozin: 70.8 ± 6.0 ng/ml vs control: 68.5 ± 10.0 ng/ml, with a difference of 2.3 ng/ml (95% CI, –6.9 – 2.3; P=.317). Other biomarkers for kidney injury, including urinary NGAL and KIM-1 in plasma and urine, did not demonstrate significant between-group differences either (Figure 4). In contrast, we did observe a difference between study groups in the development of serum HIF1α in the perioperative period. In the control group, HIF1α increased significantly from 110 ± 152 ng/ml before surgery to 171 ± 182 ng/ml on the first day after surgery (P=.04) and to 198 ± 268 ng/ml on the second day (P=.098). In contrast, in the empagliflozin group, HIF1α did not change from baseline levels in the postoperative period (Figure 4). A complete overview of all biomarker concentrations is available in Supplementary Table S1.

**Figure 4.**
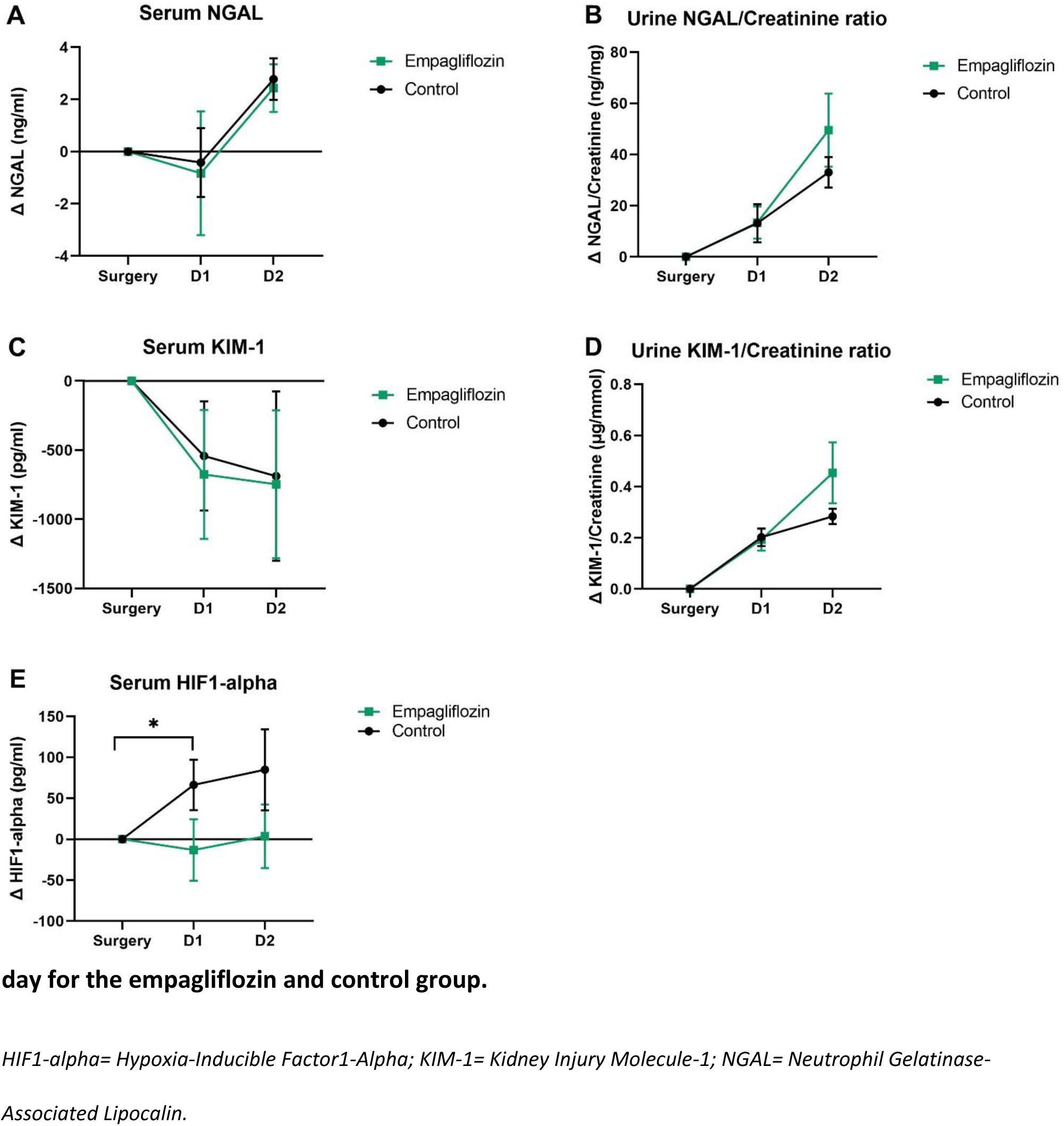
Differences in postoperative kidney biomarkers concentrations compared to surgery.

### Metabolic effects

Ketone levels increased during surgery (Figure 5) in both groups; in 41 (76%) patients, the peak levels of ketones (1.1 ± 0.6 mmol/L) were measured at the end of CPB. The increase in ketone levels did not differ significantly between the groups. In the empagliflozin group, ketone levels increased by 1.0 ± 0.5 mmol/L compared to 0.8 ± 0.5 mmol/L in the control group, difference 0.2 mmol/L (95% CI, –0.5 – 0.1; P=.208). There was no significant difference in ketone development between the groups (P value of the interaction term time*group = .75). Ketoacidosis was not observed in any of the patients.

**Figure 5.**
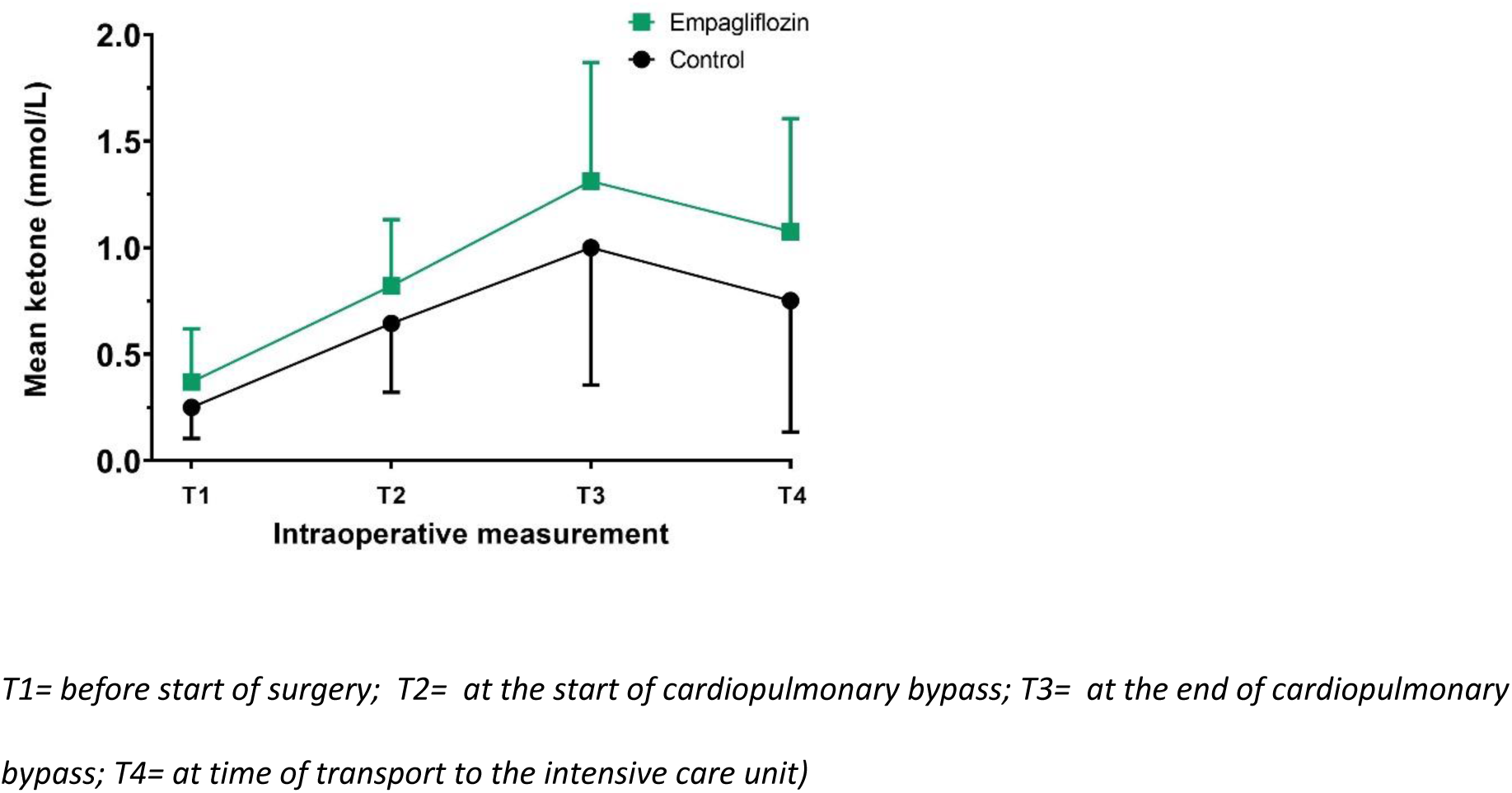
Intraoperative (mean ± SD) ketone levels.

The mean intraoperative blood glucose was 6.6 ± 1.1 mmol/L in the empagliflozin group compared to 7.1 ± 1.5 mmol/L in the control group, difference 0.5 mmol/L (95% CI, –0.3 – 1.2; P=.194). The intraoperative incidence of hyperglycemia, hypoglycemia, and insulin administrations did not differ significantly between groups (Table 2). The peak glucose concentration during the first 48 hours postoperative was 9.3 ± 1.5 mmol/L in the empagliflozin group compared to 10.9 ± 2.5 mmol/L in the control group, difference of 1.6 mmol/L (95% CI, 0.5 – 2.8; P=.007). During the postoperative period, the incidence of observed hyperglycemia was 7/25 (28%) in the empagliflozin group compared to 18/30 (60%) in the control group, difference 32% (95% CI, –56.8 – –7.2; P=.029). In the empagliflozin group, 2/25 (8%) patients required insulin administrations due to hyperglycemia, compared to 11/30 (37%) patients in the control group, with a difference of 29% between groups (95% CI, 8.4 – 48.9; P=.023). None of the participants received insulin to treat ketoacidosis.

**Table 2.** Intraoperative and postoperative glycemic control and insulin administrations.

|  | Empagliflozin<br>(n=25) | Control<br>(n=30) | Absolute<br>difference | 95% CI | P value |
| --- | --- | --- | --- | --- | --- |
| **Intraoperative** |  |  |  |  |  |
| Mean blood glucose, mean ± SD, mmol/L | 6.6 ± 1.1 | 7.1 ± 1.5 | 0.5 | -0.3 – 1.2 | 0.194 |
| Hyperglycemia (>10 mmol/L), n (%) | 3 (12) | 6 (20) | -8 | -27.2 – 11.2 | 0.487 |
| Hypoglycemia (<4 mmol/L), n (%) | 0 (0) | 1 (3) | -3.3 | -9.8 – 3.1 | 1.000 |
| Insulin administration, n (%) | 1 (4) | 2 (7) | -2.7 | -14.4 – 9.1 | 1.000 |
| **Postoperative<sup>a</sup>** |  |  |  |  |  |
| Peak blood glucose, mean ± SD, mmol/L | 9.3 ± 1.5 | 10.9 ± 2.5 | 1.6 | 0.5 – 2.8 | 0.007 |
| Hyperglycemia (>10 mmol/L), n (%) | 7 (28) | 18 (60) | -32 | -56.8 – -7.2 | 0.029 |
| Hypoglycemia (<4 mmol/L), n (%) | 0 (0) | 0 (0) | NA | NA | NA |
| Insulin administration, n (%) | 2 (8) | 11 (37) | -29 | -48.9 – -8.4 | 0.023 |

CI= confidence interval; NA= not applicable; SD= standard deviation. <sup>a</sup> The first 48 hours postoperative

### Adverse Events

No significant between-group differences were observed in the incidence of in-hospital postoperative complications (Supplemental Table 2). The length of stay in ICU was 23 (19–42) hours in the empagliflozin group compared to 32 (21–47) hours in the control group (P=.096).

## Discussion

In this open-label, randomized controlled, pilot study we investigated the effect of SGLT2 inhibition with empagliflozin on CSA-AKI. This pilot study was prematurely stopped due to the large between-group difference in AKI incidence; empagliflozin reduced the AKI incidence by 47% compared to the control group. HIF1α levels significantly increased postoperatively in the control group, whereas there was no significant change observed in the empagliflozin group. AKI reduction was not reflected in changes in NGAL and KIM-1. Ketone bodies increased significantly during surgery in both groups without an additional effect of empagliflozin. There was no significant between-group difference in the incidence of hypoglycemia (which occurred only once in the control group), while hyperglycemia was reduced.

CSA-AKI remains a frequent unresolved complication after cardiac surgery, with reported incidences varying between 20% and 70%, depending on the type of surgery, the definition of AKI used, and the protocol for AKI monitoring. (1) Our findings align with this high incidence, exemplified by an incidence of AKI in the control group of 66.7%, which can be attributed to our meticulous monitoring regimen of hourly urine output and daily creatinine measurements. This study provides compelling evidence that perioperative SGLT2 inhibition with empagliflozin offers protection against CSA-AKI. Our findings are in agreement with three meta-analyses consistently demonstrating a reduction in AKI incidence in large-scale cardiovascular and kidney outcome trials in outpatients. (10, 13, 22) The hypothesis that SGLT2 inhibitors provide kidney protection comes from the theory that SGLT2 inhibitors reduce kidney workload and may, therefore, improve kidney tissue oxygenation.

To estimate the effect of empagliflozin on kidney hypoxia responses, we measured HIF1α levels, an established biomarker for hypoxia. We found that serum HIF1α levels remained stable in the empagliflozin group, whereas HIF1α increased in the control group after surgery. This is in line with earlier studies reporting that SGLT2 inhibition suppressed the HIF1-α pathway. (23, 24) Additionally, it has been shown (in mice) that SGLT2 inhibition after ischemia/reperfusion injury decreased the level of endothelial thinning, hypoxia, and the development of kidney fibrosis.(25)

While the clinical incidence of AKI was significantly lower after SGLT2 inhibition, this was not reflected by NGAL and KIM-1 differences. We selected serum and urinary NGAL and KIM-1 because they are widely studied biomarkers for AKI. (8) This study demonstrates that NGAL and KIM-1 do not reflect the kidney protective effects of SGLT2 inhibition following cardiac surgery.

In light of the FDA warning regarding the risk of ketoacidosis in patients perioperatively treated with SGLT2 inhibitors,(26) we measured the development of ketones during surgery. We observed a rise in ketone levels during surgery; however, no significant differences were observed between treatment groups. In addition, none of the patients were diagnosed with ketoacidosis during or after surgery. It is noteworthy that, by chance, insulin-dependent T2D patients were not included in this study. Had they been included, the risk of ketoacidosis would have been mitigated through the standard administration of glucose/insulin infusion perioperatively for insulin-dependent T2D patients. Although the limited sample size cannot prove safety, these findings are reassuring for further research. We also found no between-group differences in the incidence of hypoglycemia or other adverse events, while the risk for hyperglycemia was even reduced. The lack of increased hypoglycemia fits the data from trials in patients with T2D.(27)

By design, this pilot study tested for feasibility and was limited by its sample size with a standard care, open-label control group. As such, our findings warrant confirmation in a larger placebo-controlled trial. Our group is currently leading such a large multi-center trial that will include 784 patients undergoing cardiac surgery at multiple centers within the Netherlands and is estimated to complete enrollment in 2025. (clinicaltrials.gov: NCT05590143).

## Conclusion

This pilot study indicates that SGLT2 inhibition may be a promising intervention for the reduction of CSA-AKI, requiring confirmation in a larger placebo-controlled double-blinded randomized controlled trial.

## Data Availability

Data available upon reasonable request.

## Non-standard Abbreviations and Acronyms

AKI: Acute kidney injury
CI: Confidence interval
CKD: Chronic kidney disease
CPB: Cardiopulmonary bypass
CSA-AKI: Cardiac surgery-associated acute kidney injury
eGFR: estimated glomerular filtration rate
HIF1α: Hypoxia-inducible factor 1 alpha
IU: International units
KIM-1: Kidney injury molecule 1
NGAL: Neutrophil gelatinase-associated lipocalin
KDIGO: Kidney Disease: Improving Global Outcomes
SGLT2: Sodium-glucose transporter-2
T1D: Type 1 diabetes
T2D: Type 2 diabetes

## Acknowledgements

We extend our gratitude to all the participants who generously volunteered their time and commitment to this trial. Furthermore, we want to thank all the dedicated research students who were indispensable in the process of data collection: Hoang anh Nguyen, Phebe van Straten, Mariam Zaouia, Yasmine Barkia, Gwen Blokhuis and Julika Kos.

## Details of authors’ contributions

Lars Snel: Investigation, Formal analysis, Validation, Writing – Original Draft.

Maartina Oosterom-Eijmael: Investigation, Formal Analysis, Validation, Writing – Original Draft.

Elena Rampanelli: Formal analysis, Validation, Resources.

Yugeesh Lankadeva: Writing – Review & Editing, Supervision.

Mark Plummer: Writing – Review & Editing.

Benedikt Preckel: Resources, Writing – Review & Editing, Supervision.

Daniel van Raalte: Conceptualization, Methodology, Writing – Review & Editing, Supervision.

Jeroen Hermanides: Conceptualization, Methodology, Writing – Review & Editing, Supervision.

Abraham Hulst: Conceptualization, Methodology, Writing – Review & Editing, Supervision, Funding Acquisition.

## Supplementary Tables

**Supplementary Table 1.** Kidney biomarkers were measured on surgery day and the first two postoperative days.

|  |  | **Empagliflozin** | **Control** | **Absolute** | **95% CI** | **P** |
| --- | --- | --- | --- | --- | --- | --- |
|  |  | **(n=25)** | **(n=30)** | **Difference** |  | **value** |
| **Serum NGAL, (ng/ml)** | **D0** | 68.4 ± 6.0 | 66.3 ± 10.2 | -2.1 | -6.7 – 2.6 | 0.376 |
|  | **D1** | 67.6 ± 11.4 | 65.4 ± 10.6 | -2.2 | -8.2 – 3.8 | 0.460 |
|  | **D2** | 70.8 ± 6.0 | 68.5 ± 10.0 | -2.3 | -6.9 – 2.3 | 0.317 |
| **Urine NGAL/Creatinine (ng/mg)** | **D0** | 27.6 ± 58.2 | 9.0 ± 13.1 | -18.6 | -40.9 – 3.7 | 0.100 |
|  | **D1** | 41.0 ± 74.0 | 21.8 ± 40.7 | -19.2 | -50.8 – 12.4 | 0.229 |
|  | **D2** | 77.1 ± 120.8 | 41.2 ± 33.7 | -35.9 | -82.8 – 11.0 | 0.131 |
| **Serum KIM-1 (pg/ml)** | **D0** | 1458 ± 3843 | 1848 ± 4977 | 390 | -2068 – 2849 | 0.751 |
|  | **D1** | 841 ± 1694 | 1265 ± 3072 | 423 | -636 – 1465 | 0.548 |
|  | **D2** | 712 ± 1365 | 1126 ± 2300 | 414 | -636 – 1465 | 0.432 |
| **Urine KIM-1/Creatinine (µg/mmol)** | **D0** | 0.2 ± 0.2 | 0.2 ± 0.1 | 0.0 | -0.1 – 0.1 | 0.662 |
|  | **D1** | 0.4 ± 0.3 | 0.4 ± 0.2 | 0.0 | -0.1 – 0.2 | 0.893 |
|  | **D2** | 0.7 ± 0.6 | 0.5 ± 0.2 | -0.2 | 0.4 – 0.1 | 0.199 |
| **Serum HIF1α (pg/ml)** | **D0** | 142.7 ± 235.1 | 110.0 ± 151.6 | -32.7 | -139.3 – 73.9 | 0.541 |
|  | **D1** | 129.5 ± 179.6 | 170.5 ± 182.5 | 41.0 | -57.5 – 139.4 | 0.408 |
|  | **D2** | 146.3 ± 134.7 | 198.1 ± 267.8 | 51.8 | -66.5 – 170.1 | 0.384 |

*Values are presented as mean ± SD. CI= confidence interval; D0= day of surgery; D1= first postoperative day; D2= second postoperative day; HIF1α= Hypoxia-Inducible Factor-1 alpha; KIM-1= Kidney Injury Molecule-1; NGAL= Neutrophil Gelatinase-Associated Lipocalin. SD= standard deviation*

**Supplementary Table 2.**
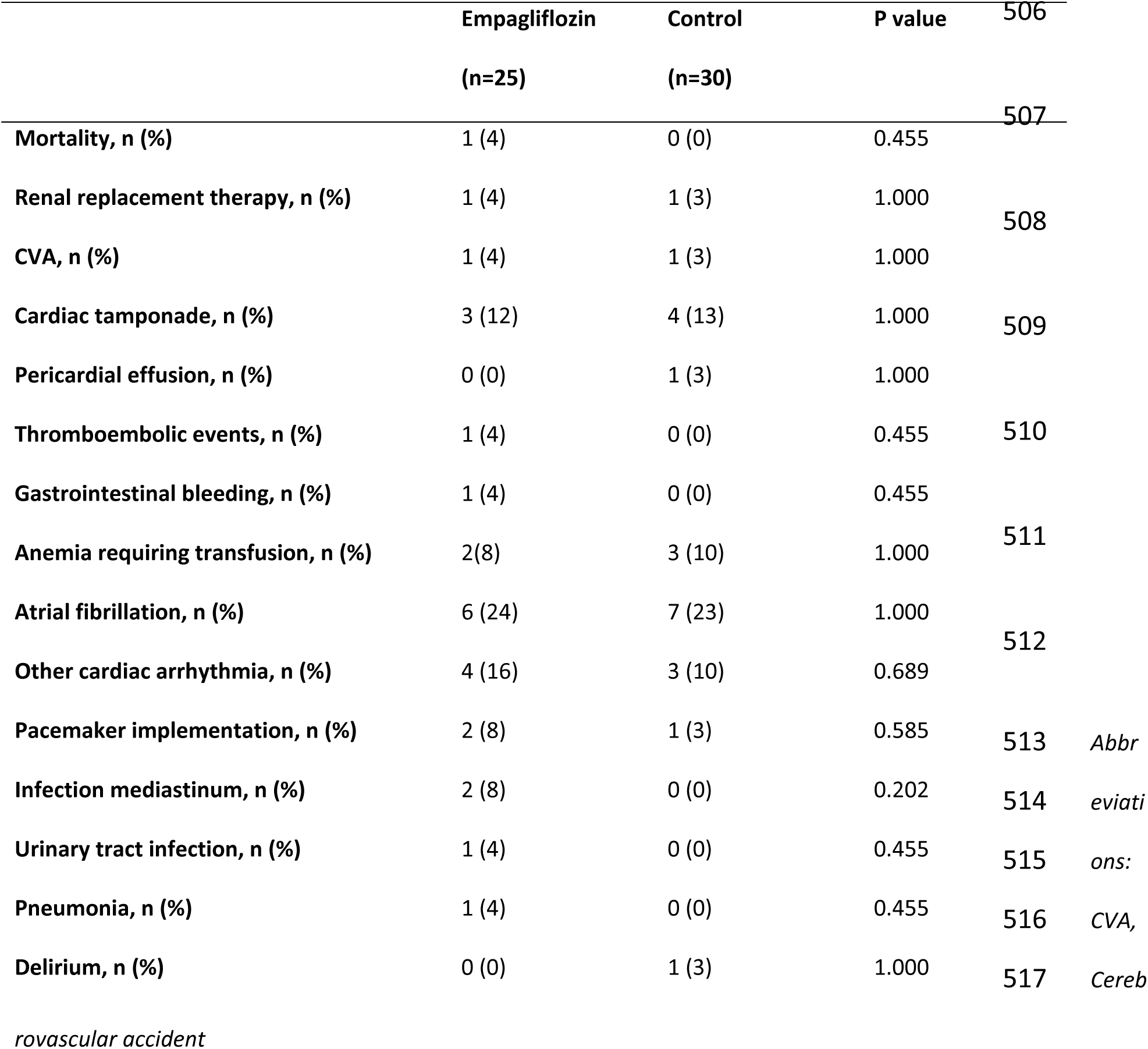
Overview of in-hospital adverse events.

|  | Empagliflozin | Control | P value | 506 |
| --- | --- | --- | --- | --- |
|  | (n=25) | (n=30) |  |  |
| Mortality, n (%) | 1 (4) | 0 (0) | 0.455 | 507 |
| Renal replacement therapy, n (%) | 1 (4) | 1 (3) | 1.000 | 508 |
| CVA, n (%) | 1 (4) | 1 (3) | 1.000 |  |
| Cardiac tamponade, n (%) | 3 (12) | 4 (13) | 1.000 | 509 |
| Pericardial effusion, n (%) | 0 (0) | 1 (3) | 1.000 |  |
| Thromboembolic events, n (%) | 1 (4) | 0 (0) | 0.455 | 510 |
| Gastrointestinal bleeding, n (%) | 1 (4) | 0 (0) | 0.455 |  |
| Anemia requiring transfusion, n (%) | 2(8) | 3 (10) | 1.000 | 511 |
| Atrial fibrillation, n (%) | 6 (24) | 7 (23) | 1.000 |  |
| Other cardiac arrhythmia, n (%) | 4 (16) | 3 (10) | 0.689 | 512 |
| Pacemaker implementation, n (%) | 2 (8) | 1 (3) | 0.585 | 513 |
| Infection mediastinum, n (%) | 2 (8) | 0 (0) | 0.202 | 514 |
| Urinary tract infection, n (%) | 1 (4) | 0 (0) | 0.455 | 515 |
| Pneumonia, n (%) | 1 (4) | 0 (0) | 0.455 | 516 |
| Delirium, n (%) | 0 (0) | 1 (3) | 1.000 | 517 |

Abbr evitati ons: CVA, Cereb
*rovascular accident*

## Supplemental Figures and Figure Legends

**Supplemental Figure 1.**
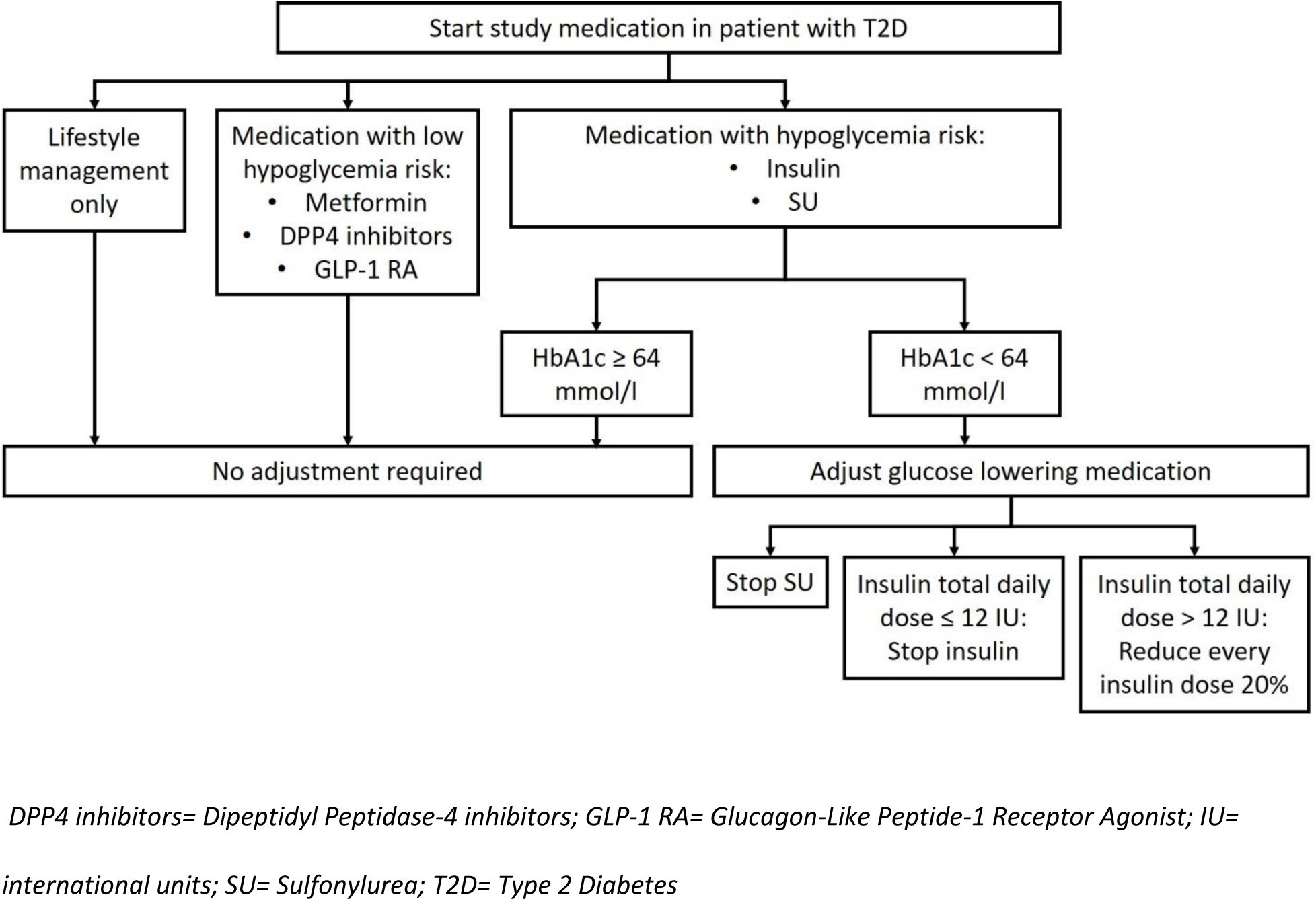
Treatment algorithm for adjustment of antihyperglycemic therapy in patients with a known history of type 2 diabetes.

